# Diagnostic Utility of Protein Biomarkers for Distinguishing Acute Ischemic Stroke and Transient Ischemic Attack: A Meta-Analysis

**DOI:** 10.1101/2025.07.09.25331200

**Authors:** Justin Y. Lin

**Affiliations:** University of California, Los Angeles, Los Angeles, CA, USA

## Abstract

With ischemic strokes being one of the most pervasive medical issues globally, identifying methods for their accurate diagnosis is becoming increasingly crucial. Two common ischemic stroke subtypes are acute ischemic stroke (AIS) and transient ischemic attack (TIA). Although these two subtypes exhibit similar symptoms, AIS is far more severe than TIA. If AIS is misdiagnosed for TIA, this can lead to inadequate treatment and worse long-term outcomes for patients. Currently, stroke diagnosis relies heavily on patient medical history and imaging techniques, causing high rates of misdiagnosis. However, identifying protein biomarker concentrations associated with AIS and TIA is a promising new method for stroke diagnosis. In this meta-analysis, ten protein biomarkers were analyzed to determine whether or not they would serve as effective diagnostic tools for AIS and TIA. After collecting the mean concentration of each biomarker from 18,160 AIS patients and 3,410 TIA patients, means were compared between AIS and TIA patient groups, determining which biomarkers had a statistically significant difference in concentration between the two stroke subtypes. Biomarkers that yielded statistically significant results were S100B, sNfl, copeptin, IL-6, and MMP-9. MMP-9 yielded the highest difference in concentration between AIS and TIA patients (536.41 ± 134.24 ng/mL in AIS patients vs. 0.11 ± 0.03 ng/mL in TIA patients). A limitation to this study was the smaller sample size of TIA patients included. While this study establishes a baseline for promising protein biomarkers that can be utilized to differentiate AIS and TIA, future studies may want to further investigate each individual biomarker, establishing clear biomarker concentration ranges for AIS and TIA.

## Introduction

Globally, ischemic strokes have become one of the most pervasive medical emergencies, with the prevalence in 2020 being 89.13 million cases (Capirossi et al., 2023). Ischemic strokes occur when a blood clot, or thrombus, blocks blood flow from an artery to the brain. Because ischemic strokes have subtypes that require different treatment methods, differentiating these subtypes during stroke diagnosis is crucial for improving patient outcomes. Two prominent types of ischemic stroke are acute ischemic stroke (AIS) and transient ischemic attack (TIA). AIS occurs when an ischemic stroke happens suddenly and is the most common stroke diagnosis (Hui, 2024). The global incidence of AIS in 2020 was 68.16 million cases, accounting for over 75% of all stroke cases that year (Capirossi et al., 2023). On the other hand, TIA occurs when there is a temporary disruption of blood flow to the brain and normally resolves on its own. In the United States, the estimated prevalence of TIA among adults is 2%. Unlike AIS, TIA results in no lasting damage to the brain and is less severe; however, TIA can serve as a warning sign of future strokes (Panuganti, 2023).

Currently, early identification is key for treating AIS, with the optimal time frame being a diagnosis within 4.5 hours of onset. While TIA can be treated using antiplatelet and anticoagulant drugs, severe cases of AIS require immediate treatments such as tissue plasminogen activator injection (tPA), intravenous thrombolysis, and endovascular thrombectomy surgery to remove blood clots (Mendelson and Prabhakaran, 2021). Still, TIA and AIS are difficult to differentiate due to the two sharing very similar symptoms. In fact, roughly 50% of patients with transient neurologic deficits have an uncertain diagnosis prior to neuroimaging (Coutts, 2017). If AIS is misdiagnosed for TIA, this can lead to inadequate treatment and worse long-term implications for stroke patients, with stroke misdiagnosis accounting for 40,000-80,000 preventable deaths annually (Newman-Toker et al., 2014).

In clinical settings, stroke diagnosis relies heavily on patient medical history, most notably information about past diagnoses (previous stroke, diabetes, heat disease, etc.). Although neuroimaging techniques like CT scans are used for stroke diagnosis, establishing other methods for accurately diagnosing stroke can improve accuracy moving forward (Dagonnier et al., 2021). Particularly, identifying protein biomarker concentrations associated with ischemic stroke subtypes is a promising new method for differentiating AIS and TIA. Although widely studied for stroke, previous clinical trials have only examined the concentration of protein biomarkers for AIS and TIA in isolation or within specific contexts, without comprehensive comparison across various studies. This meta-analysis aims to fill this gap by extracting data from past clinical trials, comparing differences in protein biomarker concentration for AIS vs. TIA patients. With this data, this study’s purpose is to identify protein biomarkers that can help clinicians effectively diagnose AIS vs TIA.

## Literature Review

According to the National Institute of Health, a biomarker is a biological molecule found in blood, bodily fluids, or tissues that signals a normal biological process, pathogenic process, or pharmacological response (Strimbel and Tavel, 2010). For strokes, elevated or depressed levels of protein biomarkers can serve as a useful tool for prehospital diagnosis, reducing the need for brain imaging and saving time that can be allocated towards treatment.

### Clinical Methodologies for Protein Biomarker Collection

In humans, protein biomarkers are collected from biological fluids such as blood and cerebrospinal fluids. First, blood samples are collected using venipuncture, and cerebrospinal fluid is collected through a lumbar puncture. Compared to cerebrospinal fluid, collecting blood samples via venipuncture is quicker and less invasive, making blood-based protein biomarkers easier to measure for strokes (Alawode et al., 2021). To prepare samples for protein analysis, biological fluid is first centrifuged to remove any debris or impurities before being stored in −80°C. Subsequently, the concentration of protein biomarkers is measured using enzyme-linked immunosorbent assays (ELISA), mass spectrometry, or single-molecule arrays (SiMoA). ELISA works by binding antibodies to a target protein biomarker, generating a color change or fluorescence proportional to the amount of protein present (Tabatabaei and Ahmed, 2022). Mass spectrometry works by ionizing protein peptides and measuring their mass-to-charge ratios, enabling accurate quantification of proteins based on known fragment patterns and intensities. Finally, SiMoA isolates and measures individual protein molecules on beads within micro-wells, allowing scientists to detect extremely low concentrations of biomarkers (Dong et al., 2023). Protein concentrations are then expressed in units such as picograms per milliliter (pg/ml), a standard unit for biomarker studies.

### Overview of Biomarkers for Ischemic Stroke

To date, over 150 candidate stroke biomarkers have been analyzed for their diagnostic potential (Dagonnier et al., 2021). For example, a study conducted by Cavrak et al. in 2021 found that AIS patients were 5.3 times more likely than TIA patients to have elevated white blood cell percentages, an immune response biomarker. MicroRNA profiling is also promising for differentiating AIS and TIA, with a 2022 study identifying 11 differentially regulated miRNAs in AIS vs. TIA patients (Toor et al., 2022). Concentrations of stroke biomarkers, most notably inflammatory biomarkers, are also higher in elderly patients above the age of 65 (Glei et al., 2011). While gene and metabolite biomarkers have been studied for stroke, this study focuses on protein biomarkers associated with ischemic stroke. In ischemic stroke patients, different concentrations of protein biomarkers can be correlated with infarct size (death of brain tissue due to loss of blood supply) and inflammation, each revealing diagnostic differences between TIA and AIS patients.

### Protein Biomarkers Assessed in Meta-Analysis

Brain natriuretic peptide (BNP) is a hormone primarily produced by the ventricles of the heart in response to increased blood pressure or fluid overload. While it is widely used to assess heart failure, BNP has also gained attention in stroke diagnostics and prognosis due to its role in atrial fibrillation and its potential link to stroke severity and outcomes. Elevated BNP levels in stroke patients can indicate concurrent cardiac strain, as strokes—especially ischemic ones—often disrupt systemic circulation. Maruyama et al. (2014) also reported that elevated BNP concentration is associated with larger infarcts, a key symptom of ischemic stroke.

S100B is a calcium-binding protein found in glial cells, specifically in oligodendrocytes and astrocytes. Elevated S100B concentrations are associated with worse neurological outcomes. When brain cells are damaged during ischemic stroke, S100B is released into the bloodstream, where elevated levels indicate blood-brain barrier disruption, glial cell activation, and inflammation. In a systematic review conducted by Rahmig et al. in 2024, researchers found five separate studies suggesting that S100B concentration may be able to distinguish AIS from stroke mimics. While S100B levels are elevated in patients post-stroke, they are also elevated in patients who have experienced traumatic brain injuries and extracranial malignancies (Dagonnier et al., 2021).

Serum neurofilament light chain (sNfl) is a biomarker for neuroaxonal damage found in cerebrospinal fluid and blood. Within neurons, sNfl is located between axons, specifically in the cytoplasm of large-caliber myelinated axons. A higher concentration of sNfl is a strong indicator of larger infarct sizes, white matter hyperintensities, and chronic cerebral small vessel disease post-stroke. Immediately following AIS, sNfl levels increase substantially. sNfl concentrations peak in AIS patients between 7 days and 3 months post-stroke, and return to baseline levels between 6 and 9 months after stroke (Holmegaard et al., 2024).

Tumor necrosis factor alpha (TNF-α) is another inflammatory biomarker produced by immune cells such as macrophages, mast cells, and lymphocytes. In the adult brain, TNF-α regulates neurotransmitter processes and is found in glia, astrocytes, and microglia. Among these processes, TNF-α also regulates the release of glutamate and can cause progenitor cell death by abruptly stopping cell division. During ischemic stroke, TNF-α stimulates tissue and leukocyte adhesion molecules, causing oxidative stress and leading to bleeding, inflammation, or thrombosis. Elevated levels of TNF-α in ischemia patients indicate worse patient outcomes (Xue et al., 2022).

Copeptin is a glycopeptide hormone released into the bloodstream by the pituitary gland in response to stress. Copeptin release is a precursor to vasopressin secretion, an antidiuretic hormone that helps regulate the body’s water and salt levels (Dobsa and Edozien, 2013). In relation to stroke, vasopressin constricts blood vessels. Elevated levels of copeptin indicate worse outcomes in stroke patients, with Perovic et al. observing in 2017 a negative correlation between copeptin levels and functional outcome in patients post-stroke. In 2015, a study consisting of 1076 TIA patients conducted by Greisenegger et al. revealed that elevated copeptin levels are indicators of recurrent stroke.

Interleukin-6 (IL-6) is a cytokine biomarker produced by immune cells such as dendritic cells, macrophages, and neutrophils. There is a strong correlation between Il-6 and CRP production, with CRP being produced as a response to increased levels of IL-6. In a meta-analysis conducted by Papadopoulos et al. in 2022, they found that higher circulating levels of IL-6 are associated with an increased risk of ischemic stroke. IL-6 levels are also associated with pathologies underlying ischemic stroke, such as atherosclerosis (buildup of fat in artery walls). Still, IL-6 levels can be influenced transiently by acute inflammatory responses like infection, something important to acknowledge for stroke diagnosis (Papadopoulos et al., 2022).

C-reactive protein (CRP) is a pentraxin protein biomarker for inflammation produced by the liver in response to inflammatory cytokines (Zhou et al., 2024). In ischemic stroke patients, past studies have shown that a higher concentration of CRP is linked to increased inflammatory responses. Elevated CRP levels are an indicator of ischemic stroke, and elevated CRP levels in TIA patients have been associated with increased risk of stroke recurrence (Mengozzi et al., 2020). Furthermore, accessing CRP as a biomarker for long-term inflammation levels post-stroke may help guide new therapies (VanGilder et al., 2014).

D-dimer is a fibrin degradation product that is released into the bloodstream when blood clots are broken down by the body through fibrinolysis. Fibrin is a protein that forms blood clots. For ischemic stroke, D-dimer serves as a key biomarker of the hemostatic system, with increased clot formation causing an increase in D-dimer production. Clinically, D-dimer tests are routinely performed to determine treatment options for ischemic stroke, such as whether or not tPA or venous thromboembolism is necessary (Ohara et al., 2019). D-dimer levels also have prognostic value, with elevated D-dimer concentration being associated with increased stroke severity and recurrence.

Matrix metalloproteinase-9 (MMP-9) is an enzyme involved in the degradation of extracellular matrix proteins in the central nervous system, playing a key role in processes such as tissue remodeling and inflammatory response. Following ischemic stroke, MMP-9 contributes to blood-brain barrier breakdown, with levels peaking 24 hours after stroke (Dagonnier et al., 2021). Higher MMP-9 concentration is also associated with the development of hemorrhagic transformation and larger infarct sizes. However, a cohort study of 255 patients conducted by Maestrini et al. in 2020 did not conclude that MMP-9 concentration is correlated with worse stroke outcomes.

Glial fibrillary acidic protein (GFAP) is an intermediate filament protein primarily expressed in astrocytes, which are critical support cells in the brain and spinal cord. GFAP is a key marker of astrocytic damage and is released into the bloodstream following injury to glial cells, making it a valuable biomarker for ischemic stroke. In clinical settings, GFAP is used to differentiate hemorrhagic and ischemic strokes, with elevated GFAP levels also being related to larger infarct sizes (Anogianakis et al., 2024).

## Materials and Methods

For the safety of patients, HIPAA regulations prevent direct access to raw patient data. Because this study requires clinical data, conducting a meta-analysis utilizing published patient data was the best course of action. Working with published data for this study was also more efficient given the range of biomarker candidates analyzed in this meta-analysis. Similar to this study, Xiang et al. in 2022 utilizes a meta-analysis to analyze cerebrospinal fluid biomarkers in patients with Parkinson’s disease.

### Study Search Strategy

First, to assess biomarker candidates for the meta-analysis, the PubMed and Google Scholar databases were searched using the following keywords: “protein biomarker AND (acute ischemic stroke OR transient ischemic attack).” Keywords are specific terms provided by an author that encapsulate the content of a research paper. “AND” ensures that both keywords are present in the papers displayed, while “OR” ensures that at least one of these keywords is present in the papers displayed. After an initial search yielding 50,226 studies, ten biomarkers present in both AIS and TIA patients were narrowed down for further analysis: BNP, S100B, sNfl, TNF-α, copeptin, IL-6, CRP, D-dimer, MMP-9, and GFAP.

Subsequently, searches specific to each biomarker and its concentration in AIS and TIA patients were performed (i.e., “acute ischemic stroke AND sNfl” and “transient ischemic attack AND sNfl”), yielding 7,220 results. Studies from these searches were then filtered on PubMed to include only cohort studies and randomized control trials, a standard procedure for collecting raw clinical data in meta-analyses. Unrelated studies not focusing on biomarker concentration were then excluded before reviewing the remaining 377 studies with a study selection criteria.

### Study Selection Criteria

The criteria for the inclusion of studies were as follows: a) the study includes the mean concentration of a target biomarker in a cohort of either AIS or TIA patients, b) the mean concentration of the target biomarker provided by the study has p ≤ 0.05, and c) the study consists of only de-identified adult stroke patients. The criteria for the exclusion of studies were as follows: a) the biomarker concentrations of patients with different categories of stroke were assessed in the same cohort, b) the study consisted primarily of pediatric patients, and c) the study did not provide the exact biomarker concentration value depicted in its figures.

Across 63 studies fitting the study selection criteria, data from 18,160 AIS patients and 3,410 TIA patients was collected. Among these studies, seven were used for collecting the mean concentrations of more than one biomarker in their respective study cohort.

### Variable Extraction

The following variables were manually extracted from the selected studies: (a) author, (b) year of publication, (c) title of study, (d) assessed biomarker, (e) mean biomarker concentration, (f) biomarker concentration p-value, and (g) sample size. Studies were first organized into ten groups based on their biomarker of interest. Within each group, studies were then split into three groups: a) studies containing biomarker concentrations of AIS patients, b) studies containing biomarker concentrations of TIA patients, and c) studies containing biomarker concentrations of separate TIA and AIS patient cohorts. Biomarker concentrations and their p-values were then extracted and organized into TIA and AIS groups.

For each biomarker’s TIA and AIS group, the mean and standard error (SEM) of the extracted concentrations were calculated using Google Sheets. Based on the metric used to measure each biomarker concentration, data was split into five CSV files.

### Meta-analysis Software

The meta-analysis was performed using RStudio Version 2025.04.0-daily+202. Data recorded on Google Sheets was imported to RStudio via CSV files, and the ggplot2 package was used to plot data comparing biomarker concentrations in AIS and TIA patients. Error bars were plotted using SEM to quantify the uncertainty of means. Overlapping error bars between AIS and TIA groups indicate a lack of statistical significance for that biomarker’s results.

## Results

### Brain Natriuretic Peptide (BNP)

For BNP, data was extracted from eight studies published between 2010 and 2017. In total, the eight studies included 2,118 AIS patients and 320 TIA patients. The mean concentration of BNP was 184.96 ± 37.80 pg/mL in AIS patients and 112.95 ± 55.03 pg/mL in TIA patients. Results for BNP were not statistically significant because of overlapping error bars (Figure 2).

**Figure 1:**
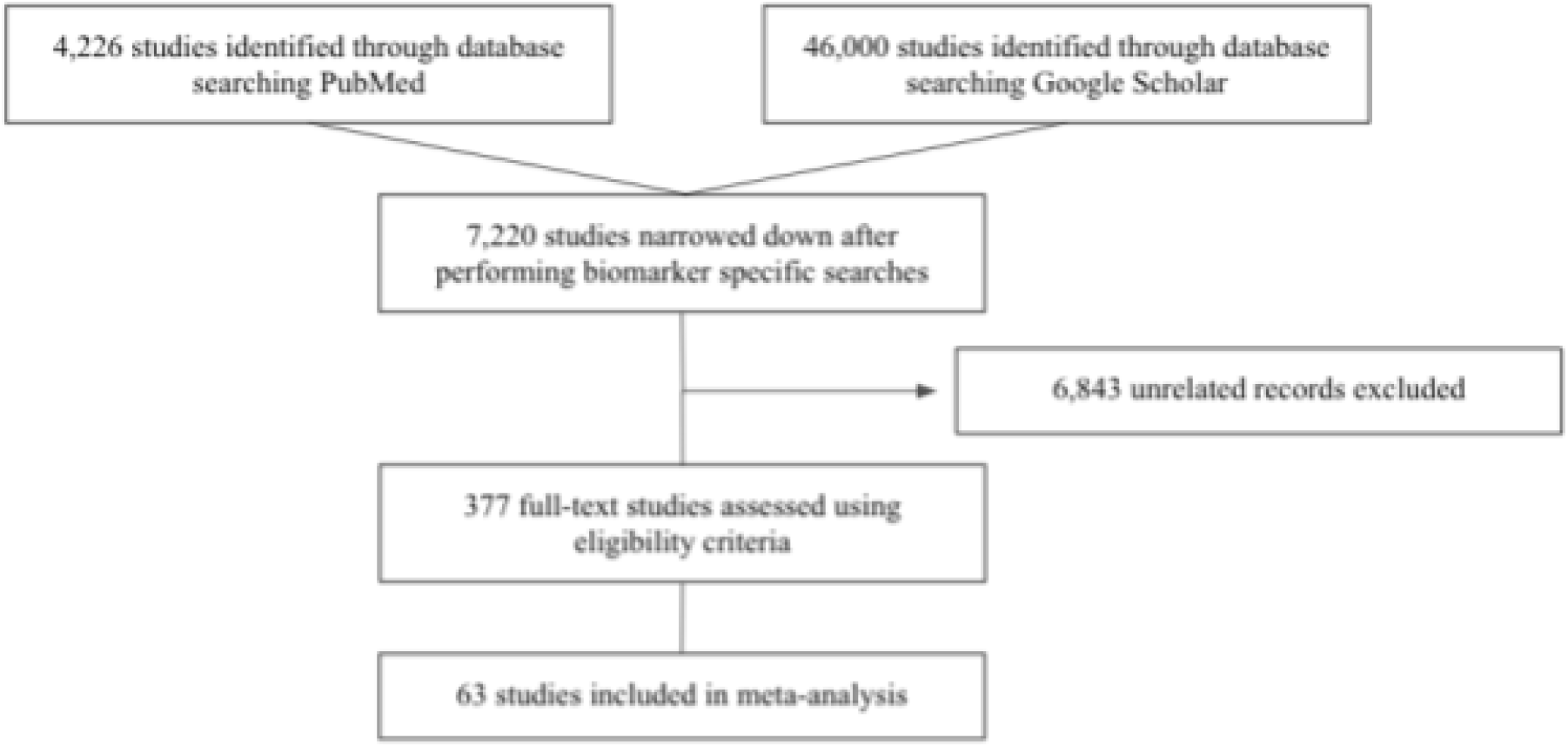
Flow chart of systematic study selection strategy.

**Figure 2:**
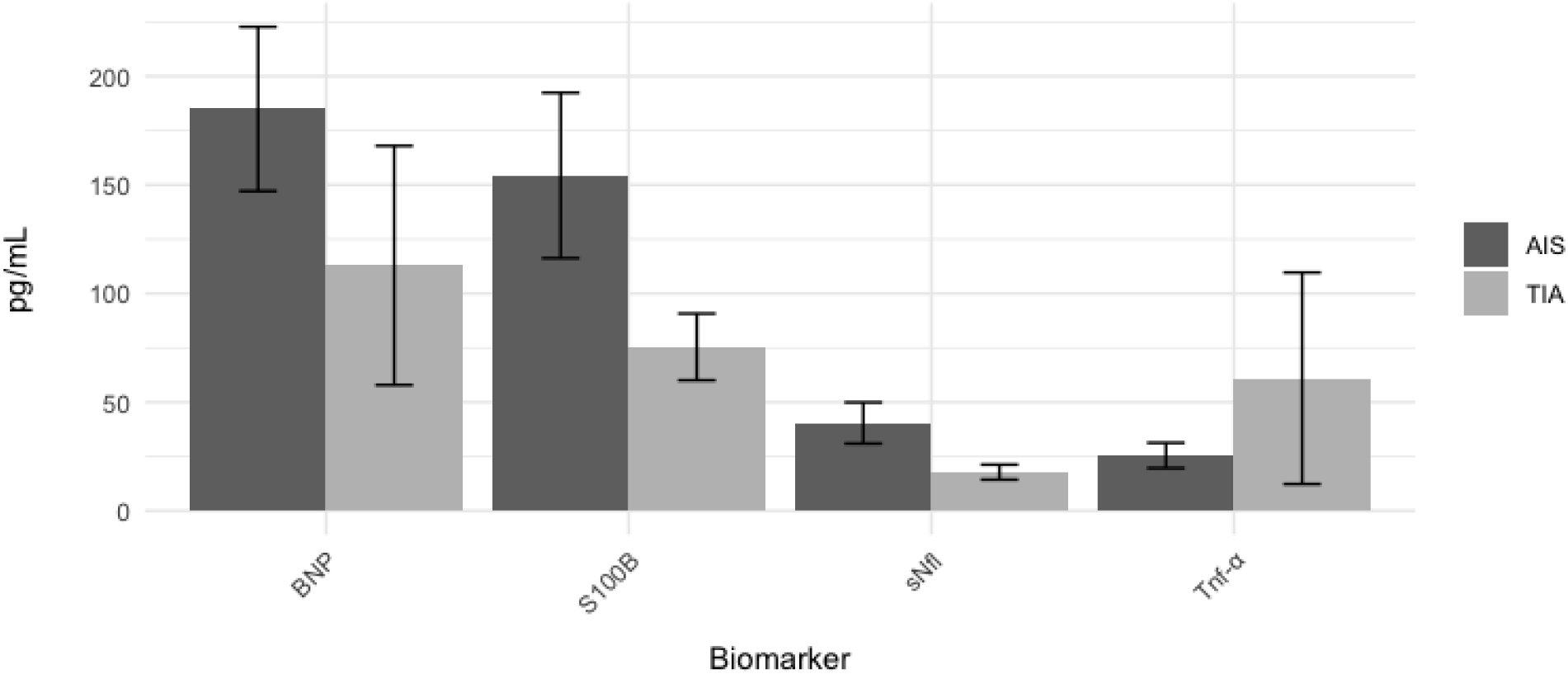
BNP, S100B, sNfl and Tnf-α Levels (pg/mL) for AIS and TIA.

### S100B

For S100B, data was extracted from five studies published between 2002 and 2020. In total, the five studies included 989 AIS patients and 55 TIA patients. The mean concentration of S100B was 154.34 ± 38.04 pg/mL in AIS patients and 75.43 ± 15.37 pg/mL in TIA patients (difference = 78.91 ± 2.40 pg/mL). Results for S100B were statistically significant (Figure 2).

### Serum Neurofilament Light Chain (sNfl)

For sNfl, data was extracted from eight studies published between 2015 and 2023. In total, the eight studies included 1658 AIS patients and 236 TIA patients. The mean concentration of sNfl was 40.41 ± 9.47 pg/mL in AIS patients and 17.83 ± 3.45 pg/mL in TIA patients (difference = 22.58 ± 0.32 pg/mL). Results for sNfl were statistically significant (Figure 2).

### Tumor Necrosis Factor Alpha (TNF-α)

For TNF-α, data was extracted from eight studies published between 2000 and 2016. In total, the eight studies included 612 AIS patients and 164 TIA patients. The mean concentration of sNfl was 25.49 ± 5.81 pg/mL in AIS patients and 61.01 ± 48.70 pg/mL in TIA patients.

Results for TNF-α were not statistically significant because of overlapping error bars (Figure 2).

### Copeptin

For copeptin, data was extracted from eight studies published between 2009 and 2016. In total, the eight studies included 1790 AIS patients and 646 TIA patients. The mean concentration of copeptin was 3.82 ± 0.24 pg/mL in AIS patients and 1.86 ± 0.42 pg/mL in TIA patients (difference = 1.96 ± 0.017 pg/mL). Results for copeptin were statistically significant (Figure 3).

**Figure 3:**
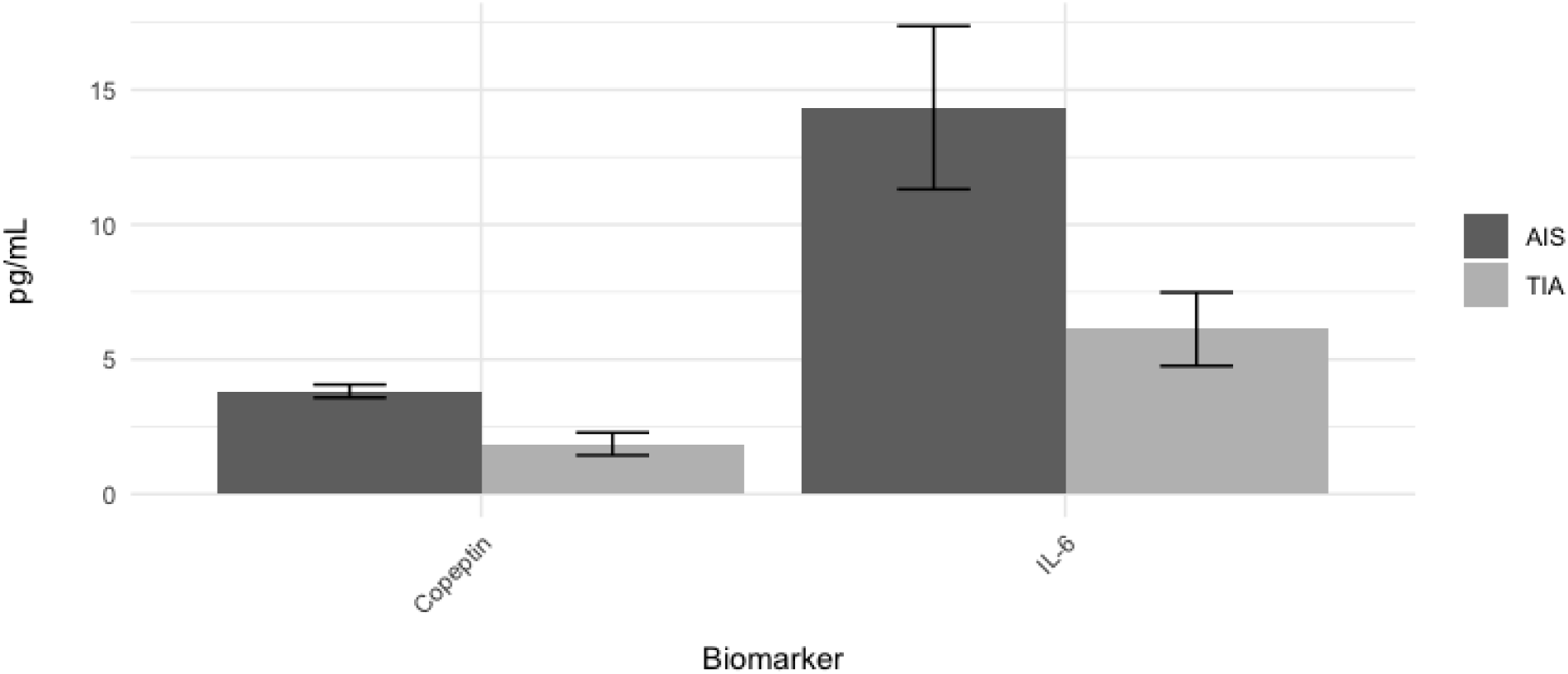
Copeptin and IL-6 Levels (pg/mL) for AIS and TIA.

### Interleukin-6 (IL-6)

For IL-6, data was extracted from nine studies published between 2000 and 2020. In total, the nine studies included 576 AIS patients and 209 TIA patients. The mean concentration of IL-6 was 14.34 ± 3.03 pg/mL in AIS patients and 6.12 ± 1.37 pg/mL in TIA patients (difference = 8.22 ± 0.16 pg/mL). Results for IL-6 were statistically significant (Figure 3).

### C-reactive Protein (CRP)

For CRP, data was extracted from six studies published between 2009 and 2020. In total, the six studies included 741 AIS patients and 348 TIA patients. The mean concentration of CRP was 4.55 ± 1.40 mg/L in AIS patients and 3.37 ± 0.90 mg/L in TIA patients. Results for CRP were not statistically significant because of overlapping error bars (Figure 4).

**Figure 4:**
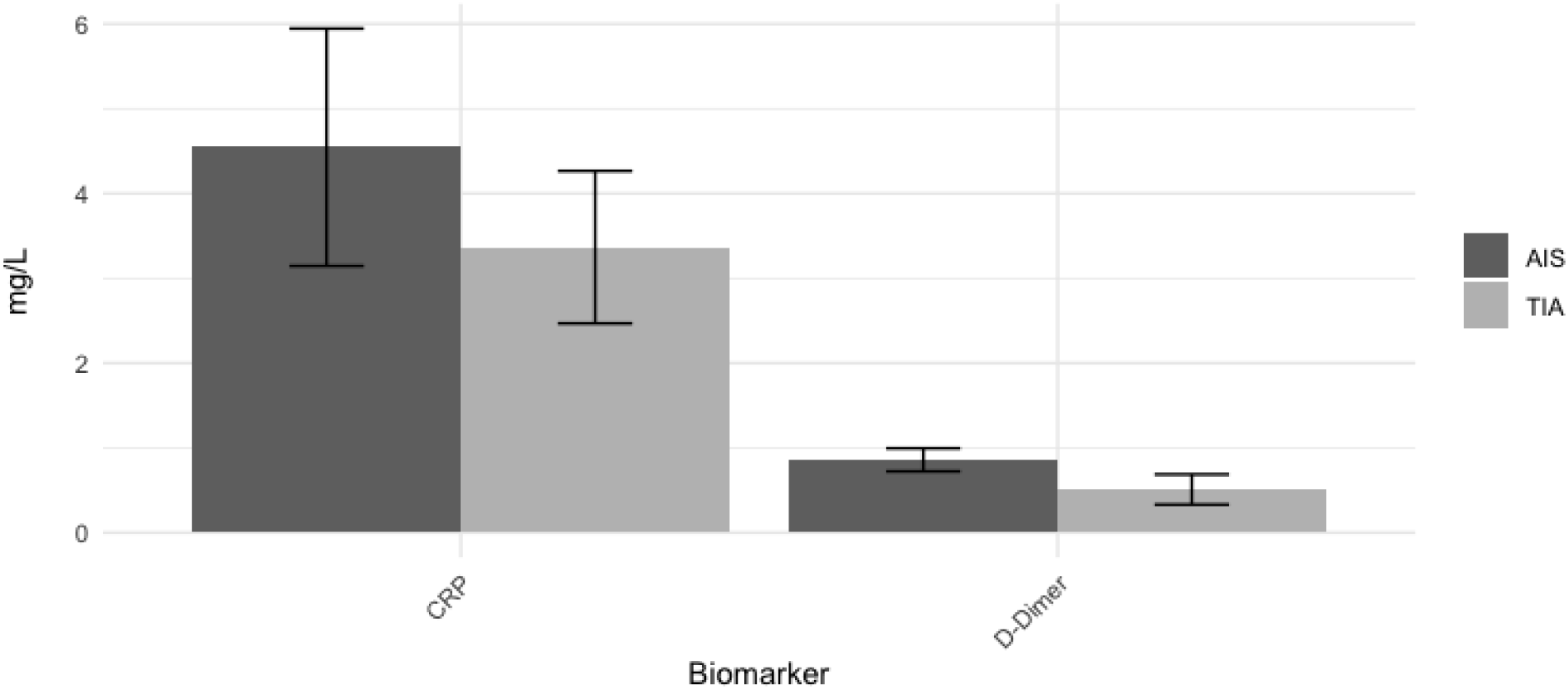
CRP and D-Dimer Levels (mg/L) for AIS and TIA.

### D-dimer

For D-dimer, data was extracted from seven studies published between 1994 and 2024. In total, the seven studies included 3,687 AIS patients and 894 TIA patients. The mean concentration of D-dimer was 0.86 ± 0.14 mg/L in AIS patients and 0.51 ± 0.18 mg/L in TIA patients. Results for D-dimer were not statistically significant because of overlapping error bars (Figure 4).

### Matrix Metalloproteinase-9 (MMP-9)

For MMP-9, data was extracted from six studies published between 2008 and 2022. In total, the six studies included 7,222 AIS patients and 842 TIA patients. The mean concentration of MMP-9 was 536.41 ± 134.24 ng/mL in AIS patients and 0.11 ± 0.03 ng/mL in TIA patients (difference = 536.3 ± 1.58 ng/mL). Results for MMP-9 were statistically significant (Figure 5).

**Figure 5:**
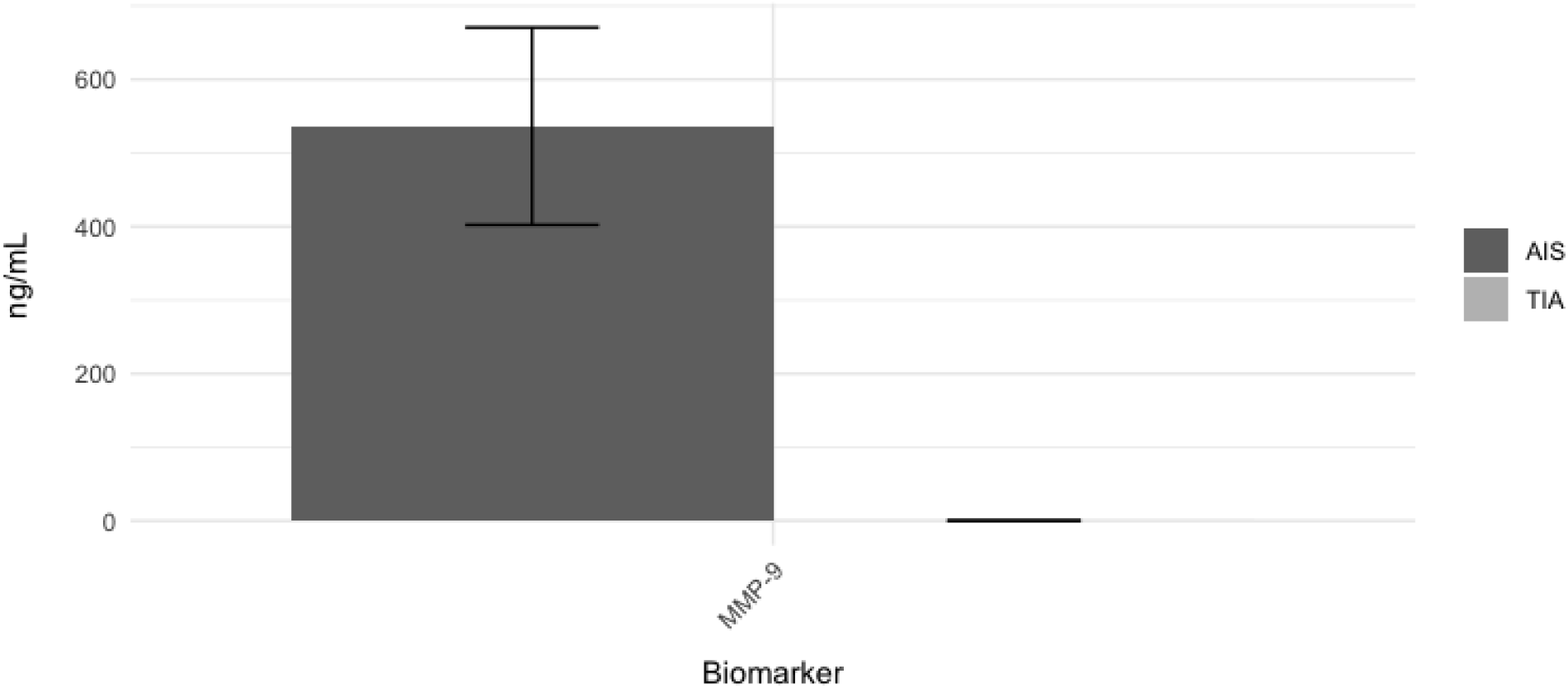
MMP-9 Levels (ng/mL) for AIS and TIA.

### Glial Fibrillary Acidic Protein (GFAP)

For GFAP, data was extracted from seven studies published between 2012 and 2023. In total, the seven studies included 930 AIS patients and 24 TIA patients. The mean concentration of GFAP was 72.17 ± 21.15 ng/mL in AIS patients and 41.7 ± 41.3 ng/mL in TIA patients.

Results for GFAP were not statistically significant because of overlapping error bars (Figure 6).

**Figure 6:**
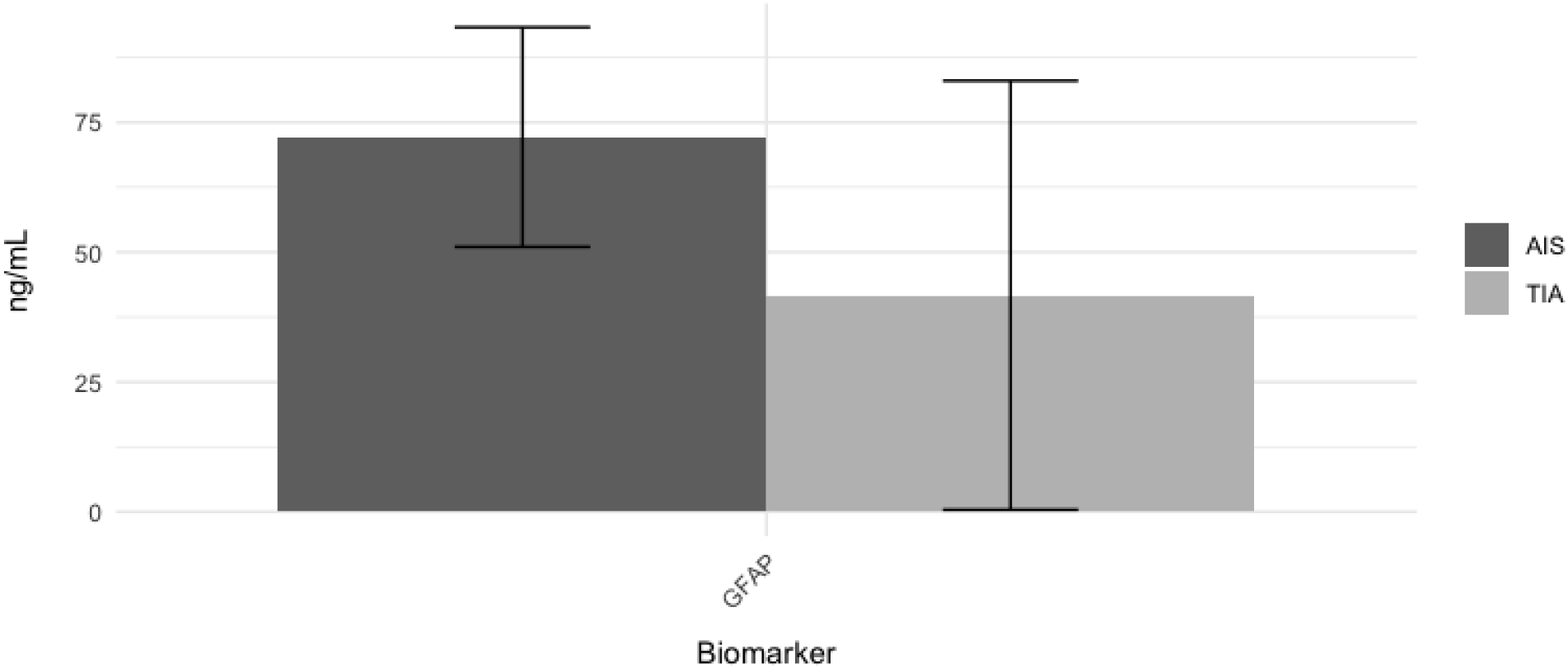
GFAP Levels (ng/mL) for AIS and TIA.

## Discussion

Incorporating clinical data from studies spanning 30 years, this study provides an updated evaluation of various protein biomarkers distinguishing AIS and TIA. Based on the results of this meta-analysis, S100B, sNfl, copeptin, IL-6, and MMP-9 have a statistically significant difference in biomarker concentration between AIS and TIA patients. For each of these five biomarkers, there is a higher concentration in AIS patients compared to TIA patients. Because a higher concentration of these biomarkers is associated with worse neurological outcomes, this pattern is reasonable, as AIS is generally more severe than TIA. The results from this meta-analysis suggest that a combination of these five biomarker concentrations may be helpful for the differential diagnosis of AIS and TIA.

Concurring with the results of Rahmig et al. in 2024, S100B serves as a promising biomarker for distinguishing AIS, TIA, and stroke mimics. Results from this meta-analysis corroborate previous studies that examine different levels of S100B expression in stroke patients, highlighting the viability of S100B as a diagnostic tool. However, it is important to recognize that S100B is also elevated in patients with traumatic brain injuries, meaning that other diagnostic factors should be used in tandem with S100B for accurate diagnoses.

Furthermore, higher concentrations of sNfl are associated with larger infarct sizes in stroke patients. With AIS patients having a higher average sNfl concentration than TIA patients, the results of this meta-analysis support previous findings indicating that AIS patients have larger infarct sizes (Ahn et al., 2022). Still, Onatsu et al. published a study in 2019 where they concluded that sNfl is correlated with larger infarct sizes, but not prognosis of AIS. Although this meta-analysis indicates that sNfl is statistically higher in AIS patients, additional studies should be conducted to verify the diagnostic utility of sNfl.

Additionally, an elevated concentration of copeptin in stroke patients is correlated with an increased risk of recurrent stroke (Greisnegger et al., 2015). Although both AIS and TIA patients have a high risk of stroke recurrence, AIS patients have a slightly higher recurrence rate (Panuganti, 2023). Because the results of this meta-analysis indicate that AIS patients have a higher concentration of copeptin, there may be a correlation between AIS, elevated copeptin levels, and a higher risk of stroke recurrence. However, it is important to note that the difference in copeptin concentration between AIS and TIA patients is much smaller compared to other biomarkers that yielded statistically significant results. Even though there is a statistically significant difference in copeptin concentration between AIS and TIA patients, this smaller difference may limit its current usage as a reliable diagnostic tool.

Next, IL-6, a protein commonly associated with inflammatory immune responses, was also found to be more elevated in AIS patients compared to TIA patients. As a biomarker, IL-6 has already been widely studied for determining the risk of stroke in patient populations.

Interestingly, IL-6 is responsible for the production of CRP, a biomarker analyzed in this study that did not yield statistically significant results. Still, expanding on past studies, findings from this meta-analysis indicate that IL-6 can potentially be utilized as a diagnostic biomarker differentiating AIS and TIA.

Most significantly, MMP-9 yielded the largest difference in biomarker concentration between AIS and TIA patients. There is a much higher concentration in AIS patients (536.41 ± 134.24 ng/mL) compared to TIA patients (0.11 ± 0.03 ng/mL). In the case of this study, strong diagnostic biomarkers should have two clear ranges of concentration: one range for AIS and one range for TIA. As a diagnostic biomarker, the results from this meta-analysis indicate that MMP-9 would be the most reliable for differentiating the two types of stroke. Because both

MMP-9 and IL-6 are associated with inflammatory responses in stroke, it would be more practical to test only one of them in a clinical setting (in this case MMP-9 since there is a much larger difference in its concentration between AIS and TIA patients). While these results do appear promising, additional studies should be conducted to verify MMP-9’s diagnostic utility.

On the other hand, biomarkers that did not yield statistically significant results include BNP, TNF-α, CRP, D-Dimer, and GFAP. With the exception of TNF-α, even biomarkers that did not yield statistically significant results had a higher concentration in AIS patients compared to TIA patients. While these results are not statistically significant, they still support a general trend: AIS patients have a higher concentration of biomarkers that correspond with worse neurological outcomes. Additional studies examining these biomarkers may help determine whether or not they can be utilized as diagnostic tools.

### Limitations

Although this study yielded statistically significant results, it is important to acknowledge key limitations. First, during the data collection phase, the number of AIS patients analyzed in this study (n = 18,160) far exceeded the number of TIA patients (n = 3,410). Each analyzed biomarker group had a higher number of AIS patients (Figure 7). Ideally, both the AIS and TIA groups should have a similarly high population size. Although the AIS group has a strong study and population size, there were far fewer studies examining biomarkers in TIA patients.

**Figure 7:**
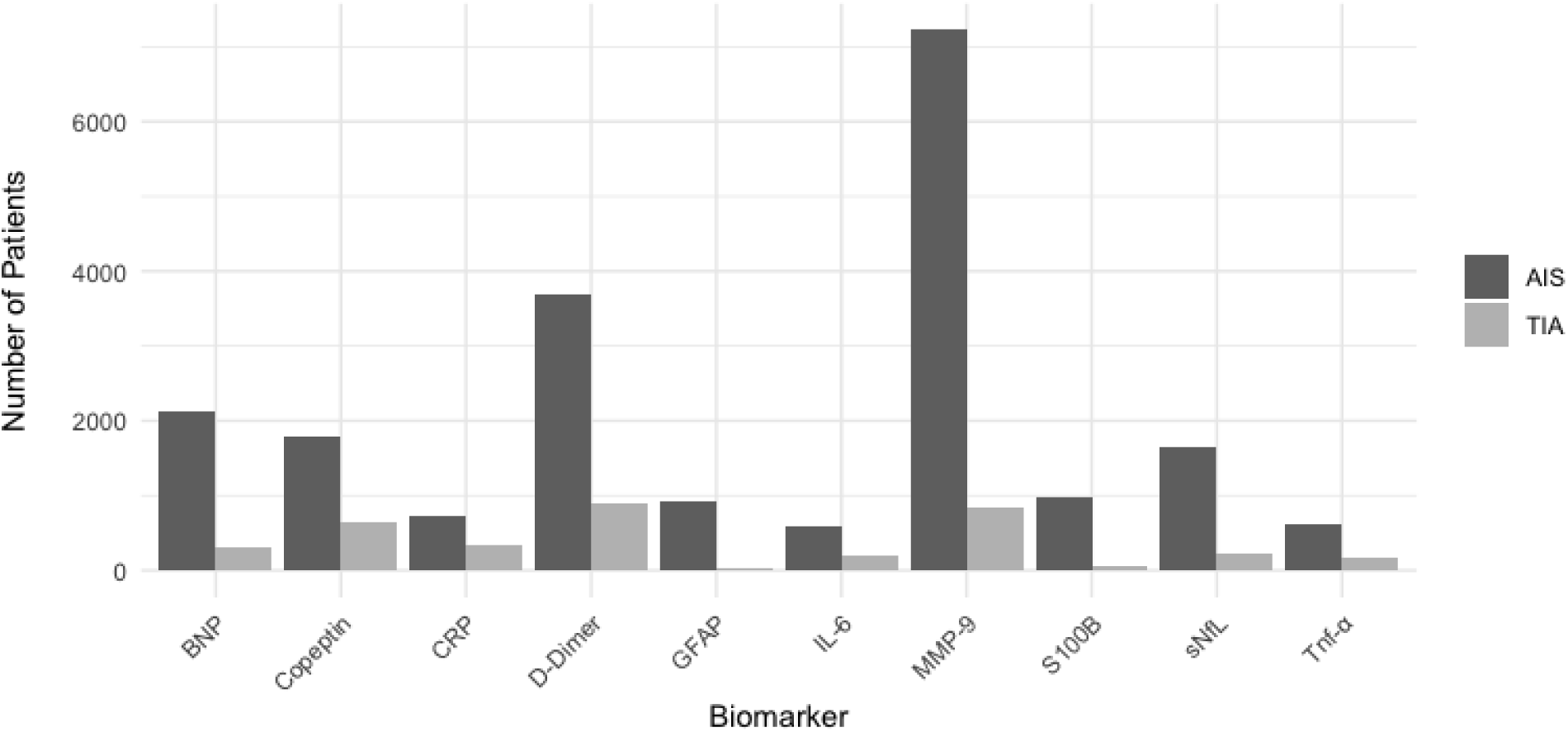
Patient Distribution of Studies Included in Meta-analysis.

Additionally, cohorts of patients analyzed in TIA studies were much smaller. This is likely due to there being fewer overall cases of TIA compared to AIS, leading to fewer studies being published about this topic. A lack of available data from TIA patients may have led to some skewed and nonsignificant results, such as TNF-α having a higher concentration in TIA patients (this result had very wide error bars). Because TNF-α is an inflammatory biomarker, it should have a higher concentration in AIS patients; however, the results from this study provided nonsignificant results showing a higher concentration in TIA patients.

In terms of the diagnostic utility of these biomarkers, it is also important to note the different metrics used when displaying concentrations. Throughout the meta-analysis, three metrics from largest to least magnitude were used: mg/L, ng/mL, and pg/mL. On figures, biomarkers are displayed in groups based on the metric that best showcases their concentration. In terms of clinically measuring these biomarkers, higher concentrations are easier to measure. Common biomarker detection methods like ELISA have a limit of detection, making concentrations at pg/mL or lower difficult to measure (Tabatabaei and Ahmed, 2022). The only statistically significant biomarker that utilizes a magnitude higher than pg/mL is MMP9, which uses ng/mL. Based on the results of this meta-analysis and MMP9’s average concentration magnitude, MMP9 is the strongest biomarker for differentiating AIS and TIA in this study.

### Conclusion

Moving forward, the findings of this meta-analysis can be utilized for future studies examining the diagnostic potential of AIS and TIA biomarkers. Due to the relative lack of TIA studies available in public databases, researchers can look into conducting additional cohort studies and randomized control trials focusing on TIA patients. For future meta-analyses, having additional TIA studies would yield results focusing on an even larger population of patients.

Furthermore, because many of these biomarkers can be affected by transient conditions not associated with stroke, biomarker concentration should not be the single determinant for diagnosing stroke. When diagnosing AIS vs. TIA, previously established tests (computed tomography scans or magnetic resonance imaging) should be performed in tandem with biomarker tests, cross-verifying the results of one another. When looking at stroke biomarkers, future studies may also want to clarify the extent to which a biomarker is affected by other conditions besides stroke. This would help narrow down the pool of eligible protein biomarkers for distinguishing AIS and TIA.

Another avenue that can be explored is establishing a clear range of biomarker concentrations associated with AIS and TIA. Overall, the main purpose of this meta-analysis was to determine which protein biomarkers have significant differences in concentration between AIS and TIA patients. This study identified five biomarkers that fit the aforementioned criteria (S100B, sNfl, copeptin, IL-6, and MMP-9), and may help researchers identify promising biomarkers for further research. To actually utilize these biomarkers as diagnostic tools, future studies should look at individual biomarkers, establishing a reliable concentration range that can be utilized to diagnose AIS or TIA. Finally, researching biomarkers besides protein biomarkers may reveal other approaches for distinguishing AIS and TIA.

## Supporting information

Supplemental Data Sheet

## Data Availability

All data produced in the present study are available upon reasonable request to the authors.

https://docs.google.com/spreadsheets/d/1SC0a9Ez3WqgStgxUOkAakpe-uzUKBy6k/edit?gid=1875644392#gid=1875644392

## Appendix

### RStudio Programming Scripts

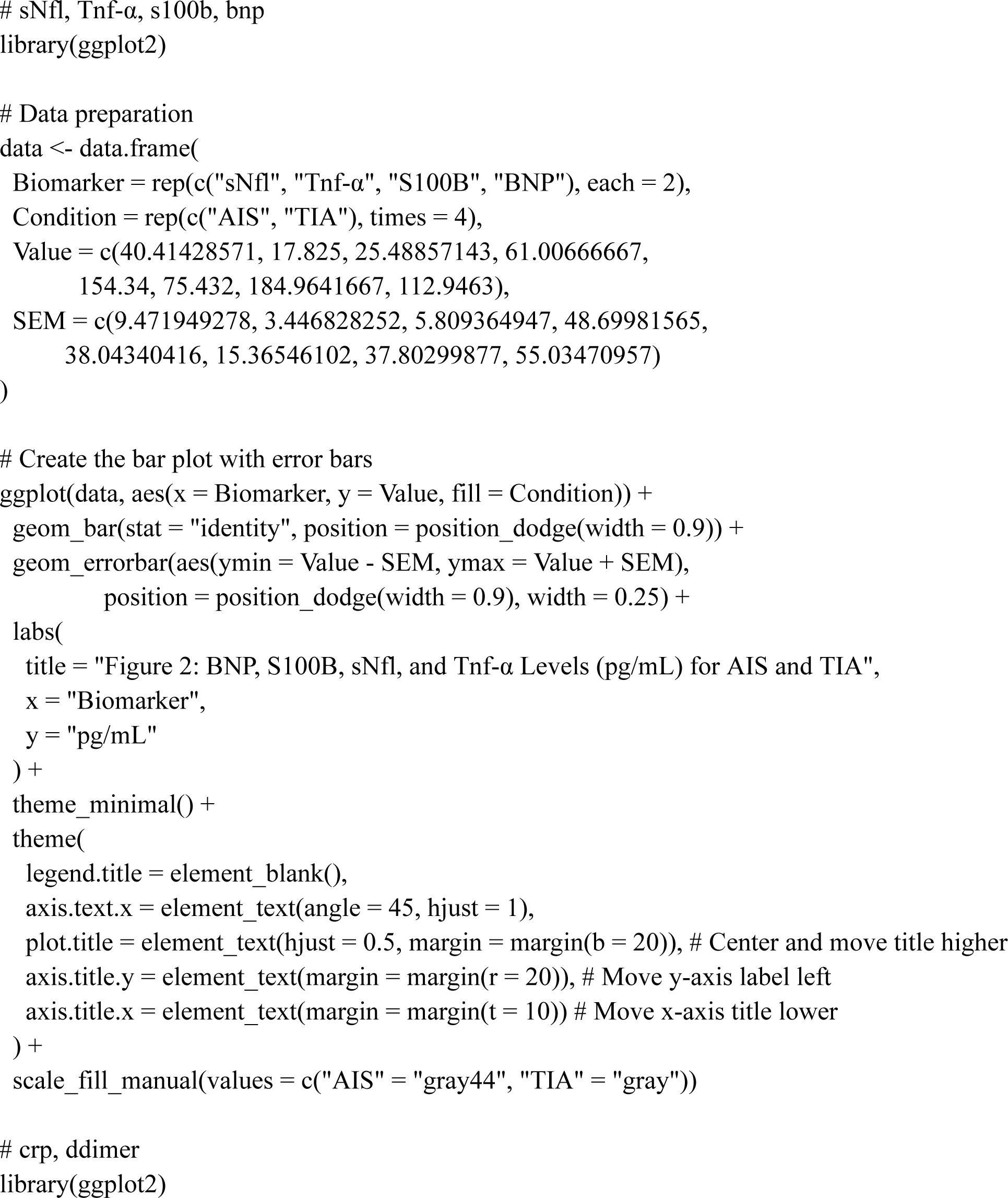
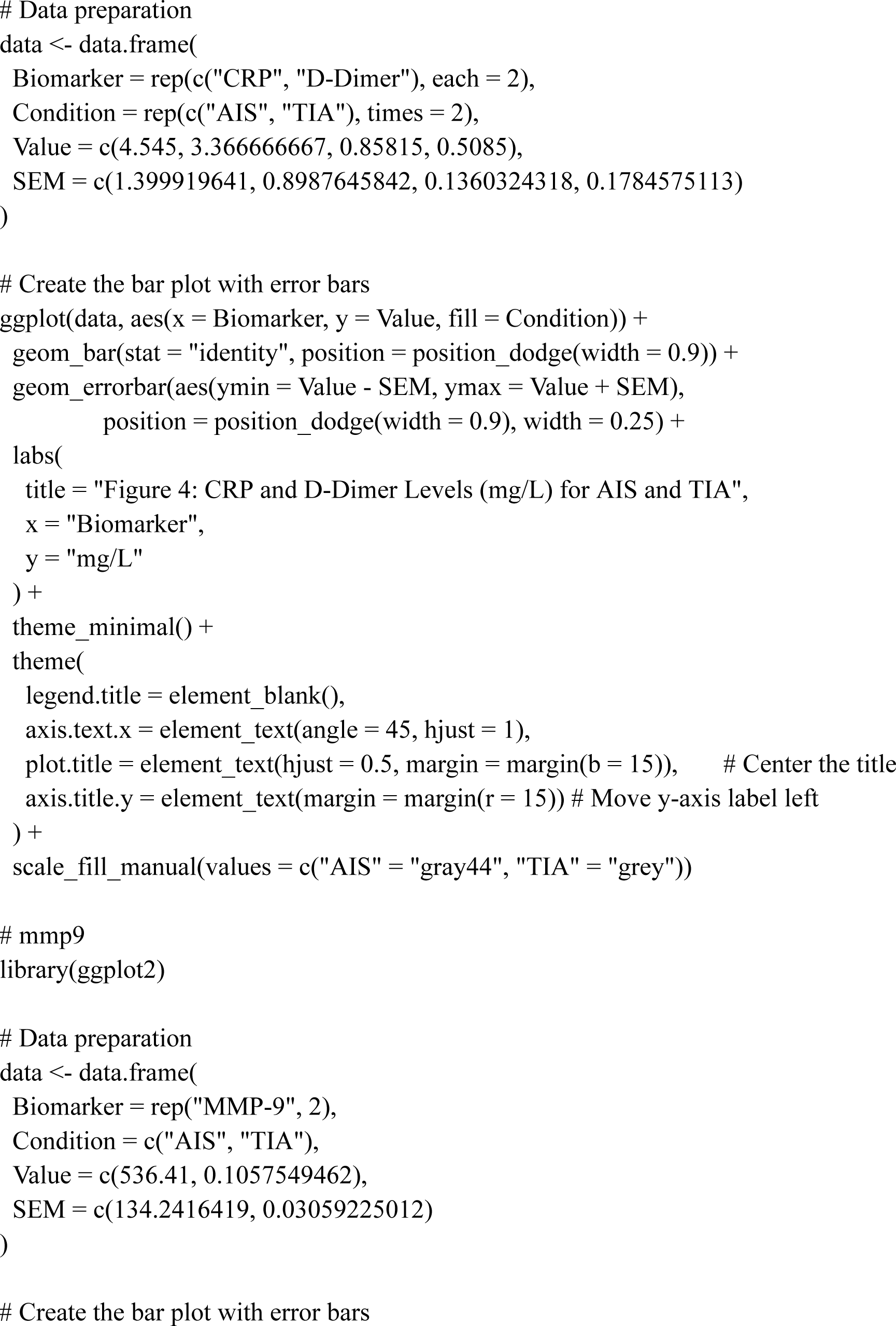
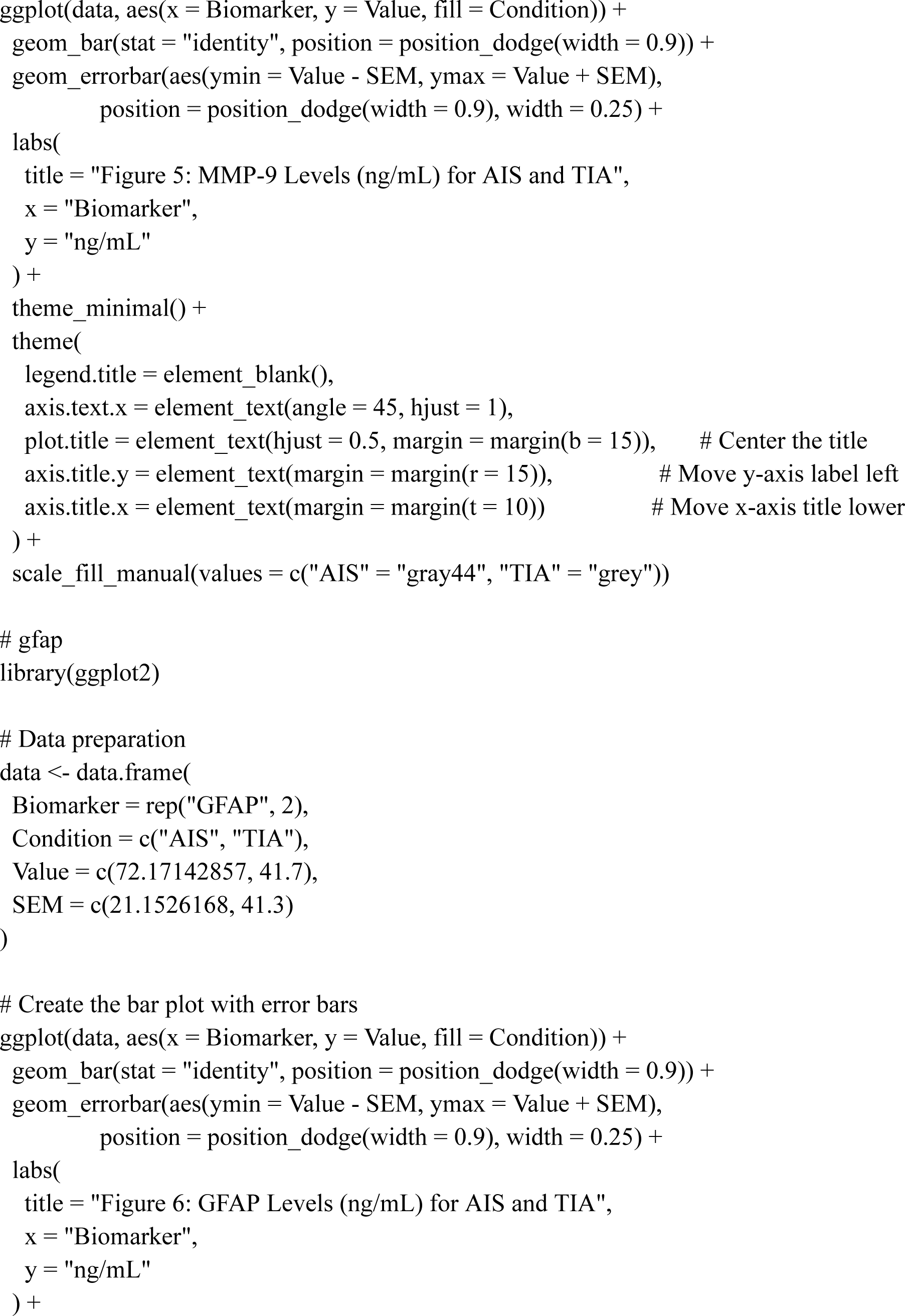
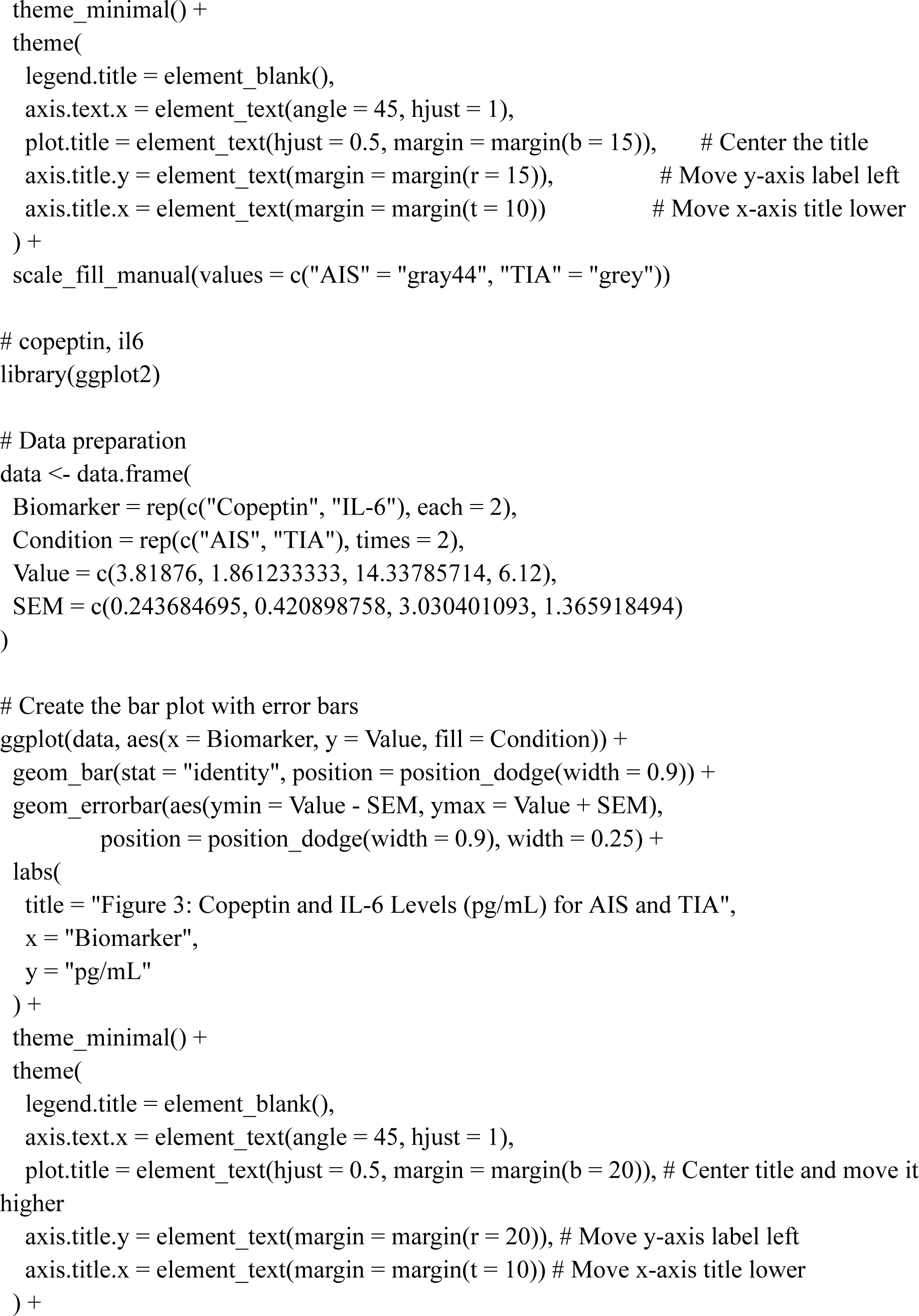
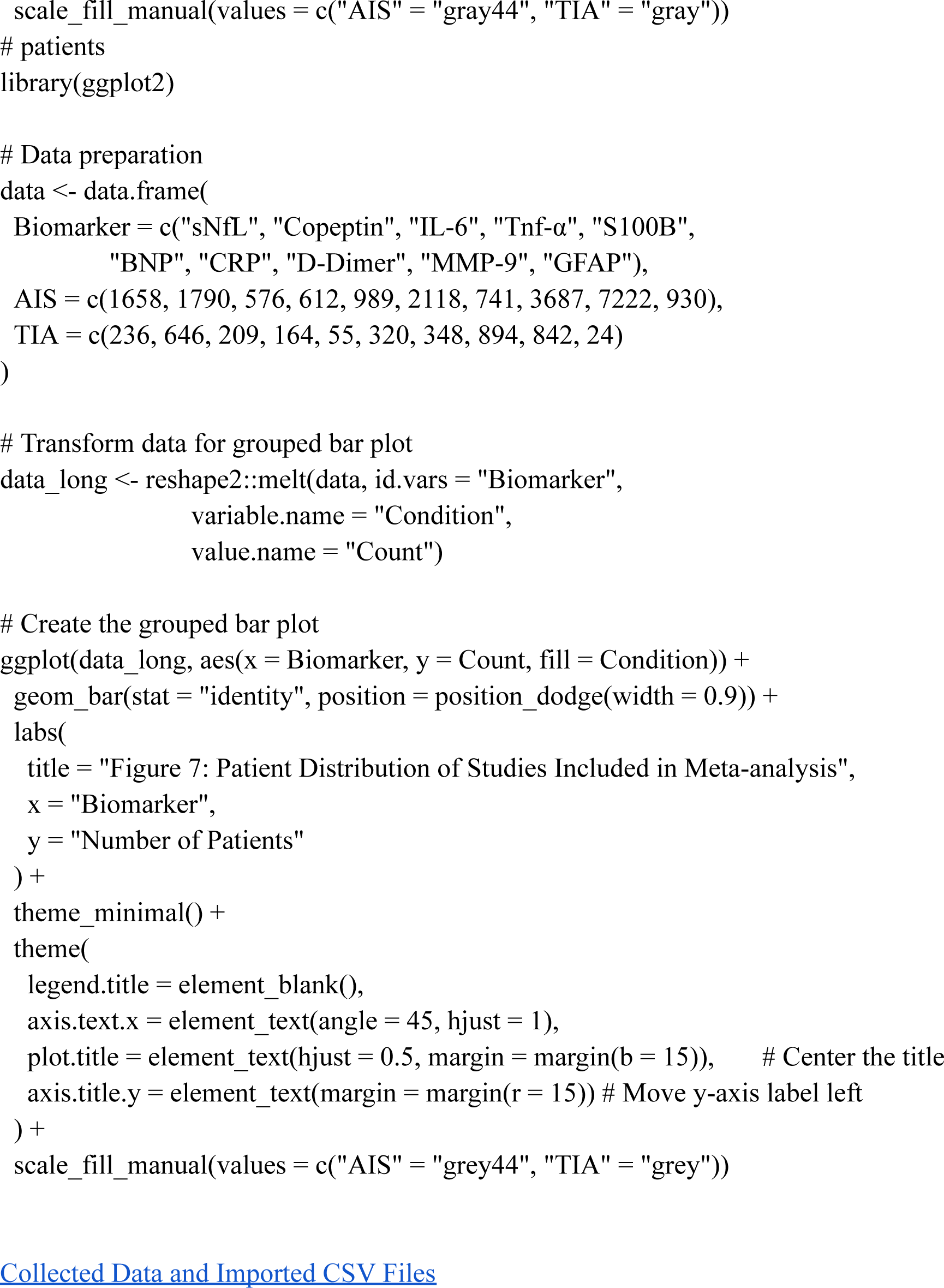

